# Quantitative chest CT combined with plasma cytokines predict outcomes in COVID-19 patients

**DOI:** 10.1101/2021.10.11.21264709

**Authors:** Guillermo Carbonell, Diane Marie Del Valle, Edgar Gonzalez-Kozlova, Brett Marinelli, Emma Klein, Maria El Homsi, Daniel Stocker, Michael Chung, Adam Bernheim, Nicole W. Simons, Jiani Xiang, Sharon Nirenberg, Patricia Kovatch, Sara Lewis, Miriam Merad, Sacha Gnjatic, Bachir Taouli

**Author notes:** These authors contributed equally (Guillermo Carbonell, Diane Marie Del Valle, Edgar Gonzalez-Kozlova). **Corresponding author:** Bachir Taouli, MD MHA, Department of Diagnostic, Molecular and Interventional Radiology, and BioMedical Engineering and Imaging Institute, Icahn School of Medicine at Mount Sinai, 1470 Madison Avenue, New York NY 10029 USA. **DATA AVAILABILITY STATEMENT** The data supporting this publication has been made available at ImmPort (https://www.immport.org) under study accession number SDY1752. The dataset has been de-identified in compliance with United States Federal Health Insurance Portability and Accountability Act (HIPAA). ImmPort is a data sharing and analysis portal for immunology research community funded by the National Institute of Allergy and Infectious Diseases (NIAID), Division of Allergy, Immunology, and Transplantation (DAIT). Please refer to the ImmPort user agreement for further details (https://www.immport.org/agreement). **CODE AVAILABILITY STATEMENT** Scripts used to query the Clarity and Caboodle databases, as well as the statistical analysis, have been uploaded to GitHub repository, https://github.com/eegk/covid19_radiology. **COMPETING INTERESTS** S.G. reports consultancy and/or advisory roles for Merck and OncoMed and research funding from Bristol-Myers Squibb, Genentech, Celgene, Janssen R&D, Takeda, and Regeneron. B.T. reports consultancy and/or advisory roles for Bayer, Helio Health and research funding/support from Bayer, Takeda, Regeneron, Echosens. **AUTHOR CONTRIBUTIONS** G.C., D.M.D.V. and E.G.K. contributed equally. S.G, S.L., M.M, and B.T., contributed to the supervision. G.C., M.C., and A.B. contributed to the radiology data curation. E.D.K, M.E., and D.S. contributed to the radiology data curation and methodology. G.C., D.M.D.V., J.X, and S.N., contributed to the clinical data curation, query design, and cohort identification. P.K. and S.N. contributed to the resources and software used to the extract data from the medical record. B.M. contributed to the formal analysis of radiology data, investigation, and methodology. G.C., D.M.D.V., E.G.K., and N.W.S. contributed to the visualizations. G.C., D.M.D.V., E.G.K., S.G., and B.T. contributed to the study concept and design, data curation, investigation, methodology, formal analysis, project administration, software, visualizations, and manuscript writing. All authors reviewed and approved final manuscript.

## Abstract

Despite extraordinary international efforts to dampen the spread and understand the mechanisms behind SARS-CoV-2 infections, accessible predictive biomarkers directly applicable in the clinic are yet to be discovered. Recent studies have revealed that diverse types of assays bear limited predictive power for COVID-19 outcomes. Here, we harness the predictive power of chest CT in combination with plasma cytokines using a machine learning approach for predicting death during hospitalization and maximum severity degree in COVID-19 patients. Patients (n=152) from the Mount Sinai Health System in New York with plasma cytokine assessment and a chest CT within 5 days from admission were included. Demographics, clinical, and laboratory variables, including plasma cytokines (IL-6, IL-8, and TNF-α) were collected from the electronic medical record. We found that chest CT combined with plasma cytokines were good predictors of death (AUC 0.78) and maximum severity (AUC 0.82), whereas CT quantitative was better at predicting severity (AUC 0.81 vs 0.70) while cytokine measurements better predicted death (AUC 0.70 vs 0.66). Finally, we provide a simple scoring system using plasma IL-6, IL-8, TNF-α, GGO to aerated lung ratio and age as novel metrics that may be used to monitor patients upon hospitalization and help physicians make critical decisions and considerations for patients at high risk of death for COVID-19.

## INTRODUCTION

Despite extraordinary international efforts to reduce the spread of SARS-CoV-2, new variants continue to appear, and outbreaks persist. Vaccine coverage remains uneven and the virus is expected to circulate throughout the global population for years to come ^1,2^. The exceptional burden placed on healthcare resources by the SARS-CoV-2 pandemic has highlighted the need for accurate predictors of disease outcomes to enable effective patient and resource management. Early identification of patients at risk for severe disease may ensure healthcare resources are properly allocated to provide maximal benefit to the population.

The World Health Organization (WHO) clinical management guidelines suggest that imaging, including computed tomography (CT), may help for diagnosis and assessment of complications in COVID-19 patients. CT imaging provides a noninvasive, rapid method for assessing COVID-19 disease severity and diagnosing complications such as pulmonary embolism ^3^. Patients infected with SARS-CoV-2 may present abnormal chest CT findings, including ground-glass opacity (GGO) and local or bilateral consolidation within 1-3 weeks of infection ^4^. Chest CT has played a critical role in the clinical care of COVID-19 patients, specifically in patient stratification and prognosis when discrepancies between clinical and chest X-rays are noted ^5^. CT qualitative scores, based on the radiologist’s evaluation of the images and CT quantitative methods, have been used to calculate pneumonia lesions burden and degree of lung involvement as potential imaging biomarkers ^6–18^. Furthermore, pneumonia lesion assessment on CT images have been shown to be predictive of severity and outcomes, potentially increasing accuracy of patient stratification ^7–12,14–16^.

The hyperinflammatory response associated with COVID-19 is known to be a major contributor of disease severity and mortality ^19^. We previously established the prognostic and predictive value of measuring cytokine levels (IL-6, IL-8, and TNF-α) in the blood of COVID-19 patients upon presentation ^20^. These cytokines are independently predictive of survival and of serious disease outcomes, including acute respiratory distress syndrome (ARDS) and multi-organ failure linked to cytokine release storms ^20,21^.

Integrating chest CT findings with clinical data and laboratory tests including CRP, procalcitonin, lymphocyte, and neutrophil counts has shown promising results in predicting COVID-19 patient outcomes ^9,13–15,18^. However, the AUC for these predictors range widely and may also be limited by biases in patient selection, model overfitting, and sometimes unclear methods ^22^.

In an effort to develop a robust tool for patient risk stratification for care prioritization, we selected maximum disease severity during hospitalization and hospital death as appropriate outcomes. We hypothesized that a combination of plasma cytokines and CT measurements would have higher predictive power of COVID-19 outcomes than either method independently. To test this hypothesis, we used our previously developed cytokine panels ^20^ in combination with CT measurements ^9–18^ to compare and evaluate the performance of predictive models for death and maximum severity. Here, we applied a data driven machine learning approach to test whether a combination of plasma cytokines and CT measurements outperform each method alone. Additionally, we also built a nomogram predicting risk of COVID-19 related death using a combination of plasma cytokine and CT variables.

## METHODS

### Cohort Design

This retrospective single-center study was approved by the Mount Sinai Health System Institution Review Board (IRB). A waiver of informed consent was obtained from the IRB to query patient’s electronic medical record (EMR). All research methods were carried out in accordance with relevant human subjects research guidelines and regulations.

Between March and September 2020, 207 patients who presented to the Emergency Departments of the Mount Sinai Health System with suspicion of COVID-19 underwent a PCR for SARS-CoV-2 infection, ELLA panel for plasma cytokines, and a chest CT to evaluate for pneumonia lesions and/or to rule out pulmonary embolism. These 207 patients were identified by querying the EPIC EMR in collaboration with the Mount Sinai Data Warehouse. Samples for the RT–PCR SARS-CoV-2 lab test were collected via nasopharyngeal or oropharyngeal swab at one of 53 different ISMMS locations, representing outpatient, urgent care, emergency, and inpatient facilities. Blood specimens for ELLA were collected via venipuncture. Chest CT scans were all performed within the Mount Sinai Health System. All specimens and imaging were collected and tested as part of standard of care.

The initial cohort of 207 patients underwent further evaluation by manual chart review. Inclusion criteria for this study were: 1) hospitalized for COVID-19, 2) plasma cytokine assessment within 48 hours upon hospital admission and, 3) chest CT up to 5 days apart from plasma cytokine assessment. We excluded patients with 1) time gap between plasma cytokine assessment and chest CT greater than 5 days (n=41), 2) CT scans with severe artifacts (n=12), 3) patients with acute conditions overlapping with COVID-19 that may have affected the cytokine assessment (n=2, a patient with acute pancreatitis, and a patient with acute cholecystitis who underwent laparoscopic cholecystectomy). Thus, our final cohort was constituted by 152 patients.

### Clinical and laboratory data

Demographic and clinical data was extracted from the Epic electronic health record (Verona, WI) for the identified patients using Epic Hyperspace (August 2019), Epic Clarity (February 2020) and Epic Caboodle (February 2020) databases via connecting to Oracle (18c Enterprise Edition Release 18.0.0.0.0) and SQL server (Microsoft SQL Server 2016 (SP2-CU11) (KB4527378) - 13.0.5598.27 (X64)) databases, respectively. Additional data elements included lab results, vital signs, need for O_2_ therapy, O_2_ saturation, chest imaging reports, clinical outcomes, and medications administered during hospitalization. Data was merged from the various data sources using R version 3.6.1 (Vienna Austria). The tables were read-in and written using R packages tidyverse (v 1.3), reshape2 (v 1.4), rms (v 6.1), glmnet (4.1), ggplot2 (v 2.0) (29) and readxl (v 1.3.1). The date of first symptoms was extracted from the electronic records via manual chart review by two independent investigators (G.C., and D.M.D.V).

### CT image acquisition

Chest CT studies were performed using a variety of vendors and systems: SOMATOM Definition AS (Siemens Healthineers, Erlangen, Germany [n=50]); SOMATOM Edge Plus (Siemens Healthineers, Erlangen, Germany [n=35]); SOMATOM Perspective (Siemens Healthineers, Erlangen, Germany[n=2]; LightSpeed VCT (GE Healthcare, Chicago, United States [n=33]); Revolution HD (GE Healthcare, Chicago, United States [n=13]); Revolution EVO (GE Healthcare, Chicago, United States [n=6]); and Aquilion Prime (Canon Medical Systems, Otawara, Japan [n=13]). A non-contrast chest CT was performed on patients with COVID-19 symptoms to evaluate for potential pneumonia lesions (n=46), and a chest CT angiogram with iodine contrast [100-200 mL of Iopamidol (Isovue, Bracco Diagnostics), depending on patient’s weight, administered by bolus injection] was performed in patients in whom pulmonary embolic disease was suspected (n=106). CT acquisition parameters are listed in Supplementary Table 1.

### CT qualitative score

Image analysis was performed by two independent experienced readers (M.C. and A.B., fellowship-trained cardio-thoracic radiologists, both with 5 years of experience) who were blinded to the clinical and laboratory data, but aware of COVID-19 diagnosis. Each reader assessed half of the cohort, with an additional overlap of 40 cases to assess inter-observer reproducibility. A CT qualitative score was calculated according to the percentage of lung parenchyma affected by GGO and/or consolidations^23,24^. Each lobe was classified as: none (0%), minimal (1-25%), mild (26-50%), moderate (51-75%), and severe (76-100%) involvement. Lung lobes with no involvement were scored as 0, minimal involvement as 1, mild involvement as 2, moderate involvement as 3, and severe involvement as 4. An overall CT qualitative score was obtained by summing the ordinal value for each of the 5 lung lobes and yielding a final score of 0-20. Additionally, the readers assessed several qualitative variables in accordance with the Fleischner society definitions^24^ (Supplementary Table 2).

### CT quantitative assessment

Using the open-source software 3D slicer *(www.slicer.org)*^25^ and the Chest Imaging Platform plug-in (*chestimagingplatform.org*), semiautomated segmentation of the lungs and intensity thresholding was performed to define the following lung regions: 1) aerated lung: <−500HU (Hounsfield Units); 2) GGOs: −500 to −100HU; and 3) consolidations: −100 to 100HU. Mediastinum, hilar structures, and pleural effusions were not included in the segmentations. All segmented images were reviewed by a single observer (G.C., a post-doctoral fellow with 8 years of experience) to evaluate the segmentation task, and manual corrections were performed if needed. A second reader (B.M., PGY-5 radiology resident) performed manual corrections on 15 patients randomly selected to assess inter-observer variability. Both observers were blinded to outcome. This quantitative analysis yielded the following variables: 1) total lung volume (mL); 2) well-aerated lung volume (mL); 3) GGO volume (mL); 4) consolidation volume (mL); and 5) GGO to aerated lung ratio.

### ELLA cytokine platform

The ELLA platform is a rapid cytokine detection system based on four parallel singleplex microfluidics ELISA assays run in triplicate within cartridges following the manufacturer’s instructions. In March 2020, as the number of COVID-19 cases was increasing in New York City, we transferred the ELLA methodology to the Clinical Laboratories at Mount Sinai Hospital, which allowed the ELLA cytokine test to be coded into our electronic health record ordering system as part of a COVID-19 diagnostic panel. This panel measures key cytokines, IL-6, IL-8 and TNF-α used for predicting patient outcomes in the setting of COVID-19^20^. We applied established cutoffs for high/low cytokine levels as follows (in pg.ml^-1^): >70 for IL-6, >50 for IL-8, and >35 for TNF-α^20^.

### Study endpoints

The endpoints were: 1) COVID-19-related death during hospitalization (hospital death) and 2) maximum severity score attained during hospitalization. We applied the WHO ordinal scale (from 0 to 7 points) to assess disease severity (prior to death)^26^.

### Statistical analysis

To test associations with outcomes, we performed Wilcoxon rank, Fisher exact, Spearman correlation^27^ and Fisher independence tests^28^ for each variable. Next, to assess the probability of survival, we performed univariate and multivariate Cox proportional hazard models for cytokines (IL-6, IL-8, TNF-α), demographic variables (age, sex, race/ethnicity), BMI, minimum O_2_ saturation upon presentation, CT qualitative score and CT quantitative variables in the coxph and survminer package on R^29^. The variables IL-6, IL-8 and TNF-α, were converted to binary variables based on thresholds previously described in the methods section^20^. The threshold used for accepting null hypothesis was set to adjusted p-value<0.05 after false discovery rate correction for multi-observation correction. Additionally, for Fisher independence test, we binned numeric variables into 4 quartiles. To further simplify the severity endpoint, we binned patients into mild (3-4) and severe (5-7) groups according to the WHO ordinal scale. Maximum severity was set as the highest degree of severity at any point during hospitalization. Severity was capped at 7, prior to death when applicable.

### Model building and prediction performance

We used elastic net regression available on glmnet package^30^ to build a predictive model of outcomes: 1) death during hospitalization and 2) maximum severity score. To validate the predictions, we used a combination of cross-validation and testing/training proportions. Briefly, we maintained the proportions of cases/controls while randomly selecting 100 times samples for each of the ratios: 9/1, 8/2, 7/3, 6/4 and 5/5 of training and testing, respectively. Next, within each randomly selected subset we tested one model for each, mixing coefficient for ridge and lasso regression (alpha values of 0 or ridge to 1 or lasso), producing 500 significant (p-value <0.05) models per scenario. Additionally, we filtered variables that had a statistically significant (p-value<0.05) effect using elastic net regression coefficient selection. Model performance was evaluated using ROC analysis^31^. The comparisons between different models’ AUCs were done using Wilcoxon rank sum test, bootstrap and DeLong statistics on default settings available in the pROC package^32^. Finally, the variables defined by the optimized model were used to build a nomogram by translating the model statistics into probabilities using the rms package^33,34^.

## RESULTS

### Cohort characteristics

We obtained health information, imaging, and laboratory results as part of standard clinical care from 152 patients with confirmed COVID-19 diagnosis as seen at the Mount Sinai Health System. The median time from hospital admission to chest CT was 0.72 days (IQR 0.0 – 1.0) and to cytokine testing was 0.28 days (IQR 0.0 – 1.1). The median time between chest CT and cytokine testing was 1.0 days (IQR 0.0 – 2.0). The hospital mortality rate for our cohort was 17.1% **(Table 1)**. Additional patient characteristics and clinical features are listed in **Table 1**. The median levels for IL-6, IL-8, and TNF-α, upon presentation, were 61 pg/mL, 35.0 pg/mL, and 18.5 pg/mL, respectively **(Table 2)**. Patients who died had a higher maximum severity score (WHO ordinal scale), and lower O_2_ saturation at presentation compared to those who survived (adjusted p-value < 0.0001) **(Figure 2 & Suppl. Table 3)**. There were no significant differences in sex (adjusted p-value = 0.91), race (adjusted p-value = 0.39), ethnicity (adjusted p-value = 0.97), or age (adjusted p-value = 0.32) between patients who died vs. those who survived (by Wilcoxon rank sum test) **(Suppl. Figure 1, Suppl. Table 3)**.

**Table 1.** Patient demographics, clinical and outcome data (n=152). Data are numbers of patients with percentages between parentheses.

|  |  |
| --- | --- |
| **Median age (IQR) - years** | 61 (48 - 71) |
| **Sex (Male)** | 90 (59.2%) |
| **Race/ethnicity** |  |
| - Hispanic | 37/152 (24.3%) |
| - African American | 35/152 (23.0%) |
| - Asian | 12/152 (8.0%) |
| - White | 35/152 (23.0%) |
| - Other | 70/152 (46.1%) |
| **Obesity (BMI ≥30)** | 51 (33.6%) |
| **Oxygen saturation at presentation** |  |
| - Normal (≥95%) | 50/152 (32.9) |
| - Abnormal (<95%) | 102/152 (67.1) |
| **Comorbidities** |  |
| - Asthma | 16/151 (10.6%) |
| - Atrial Fibrillation | 12/151 (7.94%) |
| - Cancer (active) | 26/151 (17.2%) |
| - Chronic Kidney Disease | 16/151 (10.6%) |
| - Congestive Heart Failure | 14/151 (9.27%) |
| - COPD | 13/151 (8.61%) |
| - Diabetes | 43/151 (28.5%) |
| - HIV | 6/151 (4.00%) |
| - Hypertension | 52/151 (34.4%) |
| - Obstructive Sleep Apnea | 5/151 (3.31%) |
| **Smoking** |  |
| - Current | 12 (9.09%) |
| - History | 39 (61.4%) |
| **Symptoms** |  |
| - Anosmia/Ageusia | 1/152 (0.658%) |
| - Congestion/Runny Nose | 10/152 (6.58%) |
| - Cough | 78/152 (51.3%) |
| - Diarrhea | 26/152 (17.1%) |
| - Fatigue | 49/152 (32.2%) |
| - Fever | 83/152 (55%) |
| - Headache | 9/152 (5.92%) |
| - Myalgias | 20/152 (13.2%) |
| - Nausea/Vomiting | 42/152 (27.6%) |
| - Shortness of breath | 105/152 (69.1%) |
| - Sore throat | 4/152 (2.63%) |
| **Worst WHO score achieved (capped at 7)** |  |
| - Mild (3-4) | 78/150 (52.0%) |
| - Severe (5-7) | 72/150 (48%) |
| **Clinical Outcomes** |  |
| - ICU admission | 43/152 (65.4%) |
| - Acute Respiratory Distress Syndrome | 10/151 (15.1%) |
| - Died during hospital admission | 26/152 (17.1%) |

**Table 2.** Cytokine assessment, CT qualitative score and CT quantitative analysis. Data are medians with interquartile ranges (IQR) in parentheses. GGO: ground-glass opacities.

|  |  |
| --- | --- |
| **Cytokine assessment** |  |
| IL-6 (pg/mL) | 61.0 (22.8-146.3) |
| IL-8 (pg/mL) | 35.0 (20.0-59.9) |
| TNF-α (pg/mL) | 18.5 (13.0-27.4) |
| **CT qualitative score** | 9 (5-14) |
| **CT quantitative analysis** |  |
| Total lung volume (mL) | 2713 (2081-3505) |
| Well aerated lung volume (mL) | 1982 (1420-2903) |
| GGO volume (mL) | 344.5 (208.2-509.6) |
| Consolidation volume (mL) | 98.20 (49.68-252.6) |
| GGO/well-aerated lung ratio | 0.1774 (0.0767-0.3673) |

**Figure 1.**
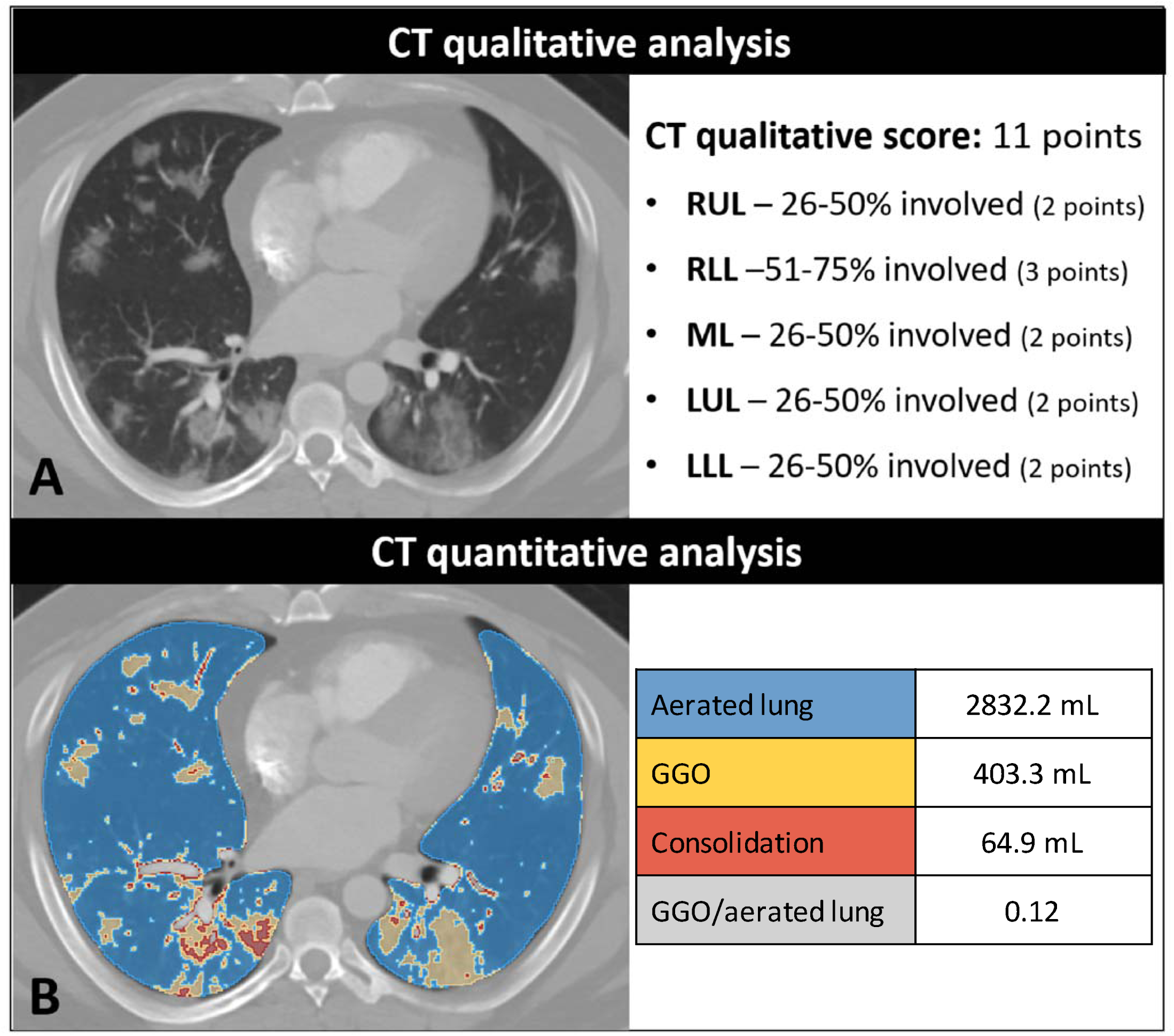
CT qualitative score and CT quantitative analysis. Male patient with COVID-19. A) CT demonstrates multifocal ground-glass opacities and regions of consolidation in the right lower. The qualitative score established by a radiologist is based on the percentage of lung involvement per lobe (shown on the right, range 0-20). B) CT quantitative analysis using segmentation software. Quantitative analysis extracts volumetric measurements (shown on the right) representing the aerated lung, the ground glass opacities (GGO) volume, the consolidation volume and the GGO to aerated lung ratio. RUL: right upper lobe; RLL: right lower lobe; ML: middle lobe; LUL: left upper lobe; LLL: left lower lobe.

**Figure 2.**
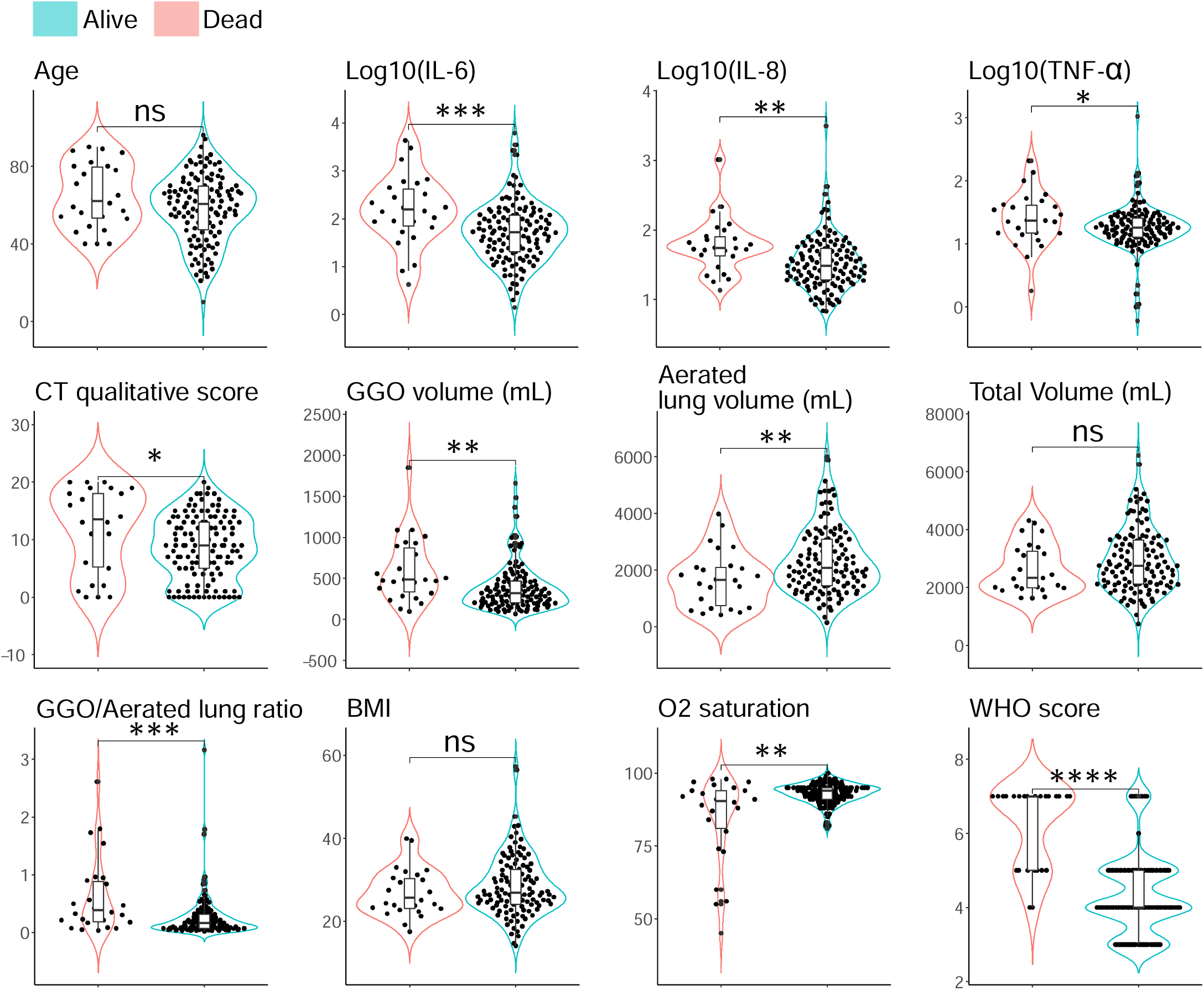
Key differences between patients who died and patients who survived. We conducted a Wilcoxon rank test for the variables collected. The number of * indicates significance (*<0.05. **<0.01, ***<0.001, ****<0.0001).

### CT qualitative score and CT quantitative analysis

CT qualitative scores were higher in patients who died vs. those who survived, but this difference did not reach significance after multiple observation correction (adjusted p-value = 0.07) **(Figure 2 & Suppl. Table 3)**. CT quantitative variables: GGO volume (adjusted p-value = 0.01), consolidation volume (adjusted p-value = 0.01), and GGO to aerated lung ratio (adjusted p-value = 0.005) were all significantly higher **(Figure 2 and Suppl. Table 3)**, while well-aerated lung volume was significantly lower (adjusted p-value = 0.035) in patients who died. A representative example of CT qualitative score and CT quantitative assessment is shown in **Figure 1**. Both CT qualitative score and CT quantitative analysis had excellent inter-observer reproducibility, with ICC of 0.998 (95% confidence interval: 0.996-0.999) and 0.986 (95% confidence interval: 0.960-0.995), respectively. **Additionally**, CT measurements and features were significantly correlated to each other, and to severity (Rho values between 0.4 and 0.7, adjusted p-values<0.05).

### Cytokine analysis

IL-6 (adjusted p-value = 0.005) and IL-8 (adjusted p-value = 0.006) levels were significantly higher in patients who died, while there was no significant difference for TNF-α (adjusted p-value = 0.119) **(Figure 2 and Suppl. Table 3)**. Also, Cytokines (IL-6, TNF-α, IL-8) were correlated to severity (Rho values between 0.3 and 0.6, adjusted p-values<0.05).

### Association between variables and risk of death

We investigate the association between a subset of clinically relevant variables and the probability of death using univariate and multivariate Cox proportional hazard models for all the available variables **(Suppl. Figures 2-10 Suppl. Tables 4-5)**. Significantly different survival probabilities were found for O_2_ saturation, IL-6, IL-8, TNF-a, Neutrophiles, Monocytes, D-dimer, and GGO to aerated lung ratio, among others (**Suppl. Tables 4, Suppl. Figures 2-10)**. Further, we observed AUCs < 0.6 (adjusted p-value < 0.05) for O_2_ saturation, age, gender, BMI and race/ethnicity **(Suppl. Table 6)**. These results indicate that oxygen saturation and demographic variables had poor power in predicting death despite producing significant prognostic models in assessing risk of death.

### Prediction of COVID-19 death

To test the hypothesis of whether cytokines or CT measurements were predictive of clinical outcomes, we developed four “scenarios” to cover all variables within a category (Cytokines or CTs). The variables selection for each of these scenarios was informed by clinical expertise, previous research, and data availability. The variables in each scenario were: 1) “Cytokines” (IL-6, IL-8, TNF-α and age), 2) “CT-Qualitative” (CT qualitative score and age), 3) “CT-Quantitative” (GGO volume (mL), well aerated lung volume (mL), Total volume (mL) and age), 4) “Combined” (All features from Scenarios 1, 2 and 3). The workflow chart highlights the steps in the construction of the predictive models is showed in **Figure 3**.

**Figure 3.**
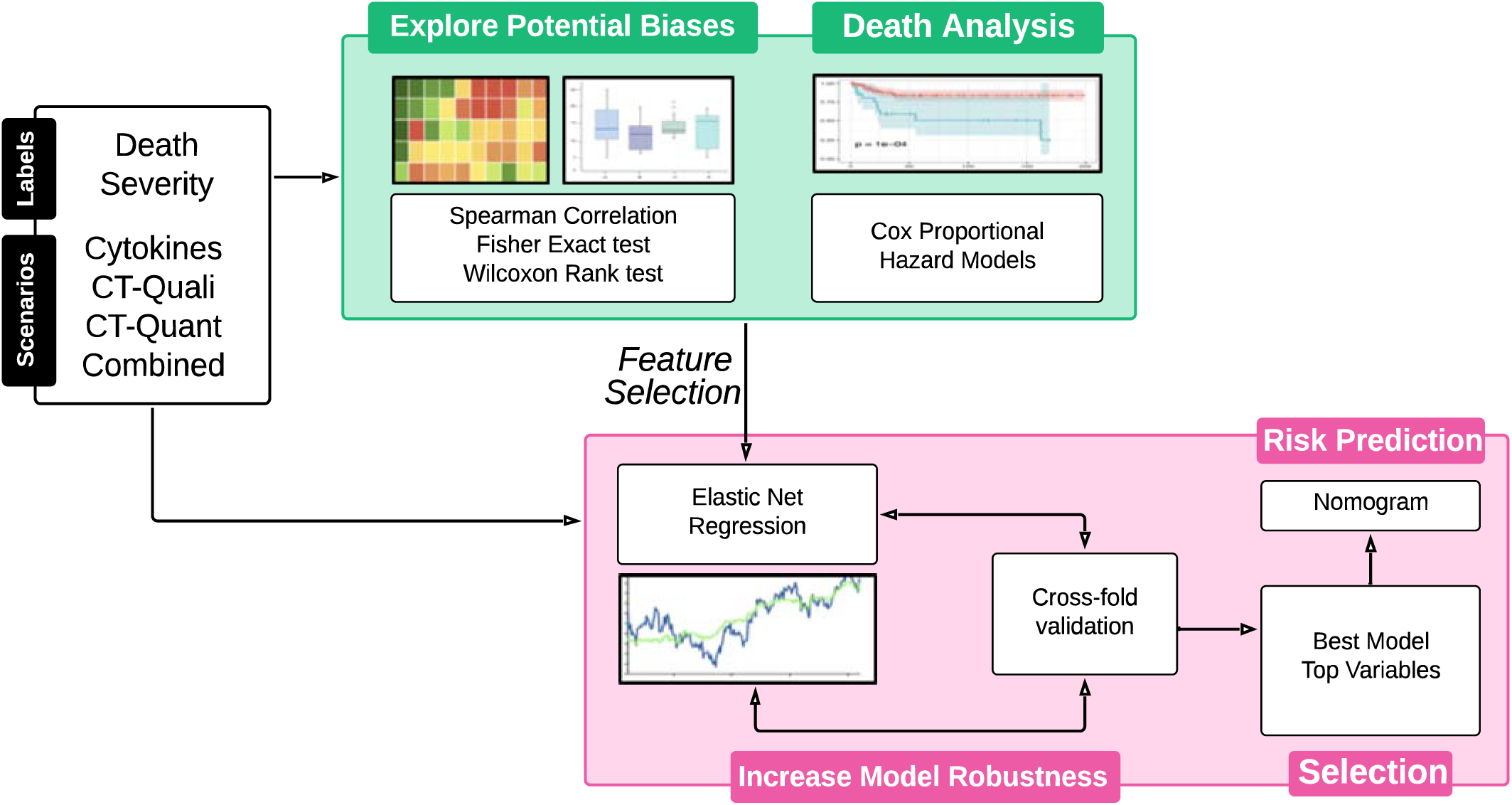
Data analysis overview. The flowchart shows how we start from 4 different scenarios (cytokines, CT qualitative, CT quantitative, combined) with the endpoints of death and maximum severity. First, we explore potential biases in our dataset by testing with several statistical approaches such as correlations, Fisher exact and independence, and Wilcoxon rank test. Next, we evaluate the probability of survival using Cox proportional hazard models to identify potential markers. Then, we use elastic net regression to explore further the predictive capabilities to separate patients that survive or not per scenario. To ensure we correctly test our hypothesis per scenario, we perform a combination of random testing/training sets and cross-fold validation to identify the predictive value of each scenario. Then, we use a coefficient-based selection to filter the significant (adjusted p-value<0.05) models and select the variables relevant for predicting. Finally, we use the variables from our best minimal model to build a risk prediction nomogram.

In parallel, to identify the best predictors of outcomes, we developed a fifth “Optimized” scenario with the aim of selecting the minimum number of variables. Thus, we used all available variables and performed a stepwise coefficient selection to choose the variables that had the highest effect on the model. Age was the only demographic variable that survived the coefficient selection process whereas race/ethnicity, BMI and sex were not selected as they did not have a significant effect on the predictive model (**Figure 3**).

The models built with cytokines showed an average AUC of 0.70 with CI 95% (65-75), while CT qualitative, CT quantitative and combined models had average AUCs of 0.61 with CI 95% (57-62), 0.66 with CI 95% (60-70) and 0.75 with CI 95% (69-80), respectively (**Figure 4A, 4B**). All models were significantly (adjusted p-values<0.05) different from each other (**Figure 4B**). These results show that a combined model increases the predictive power of death prediction of CT based on additional information from cytokine assays.

**Figure 4.**
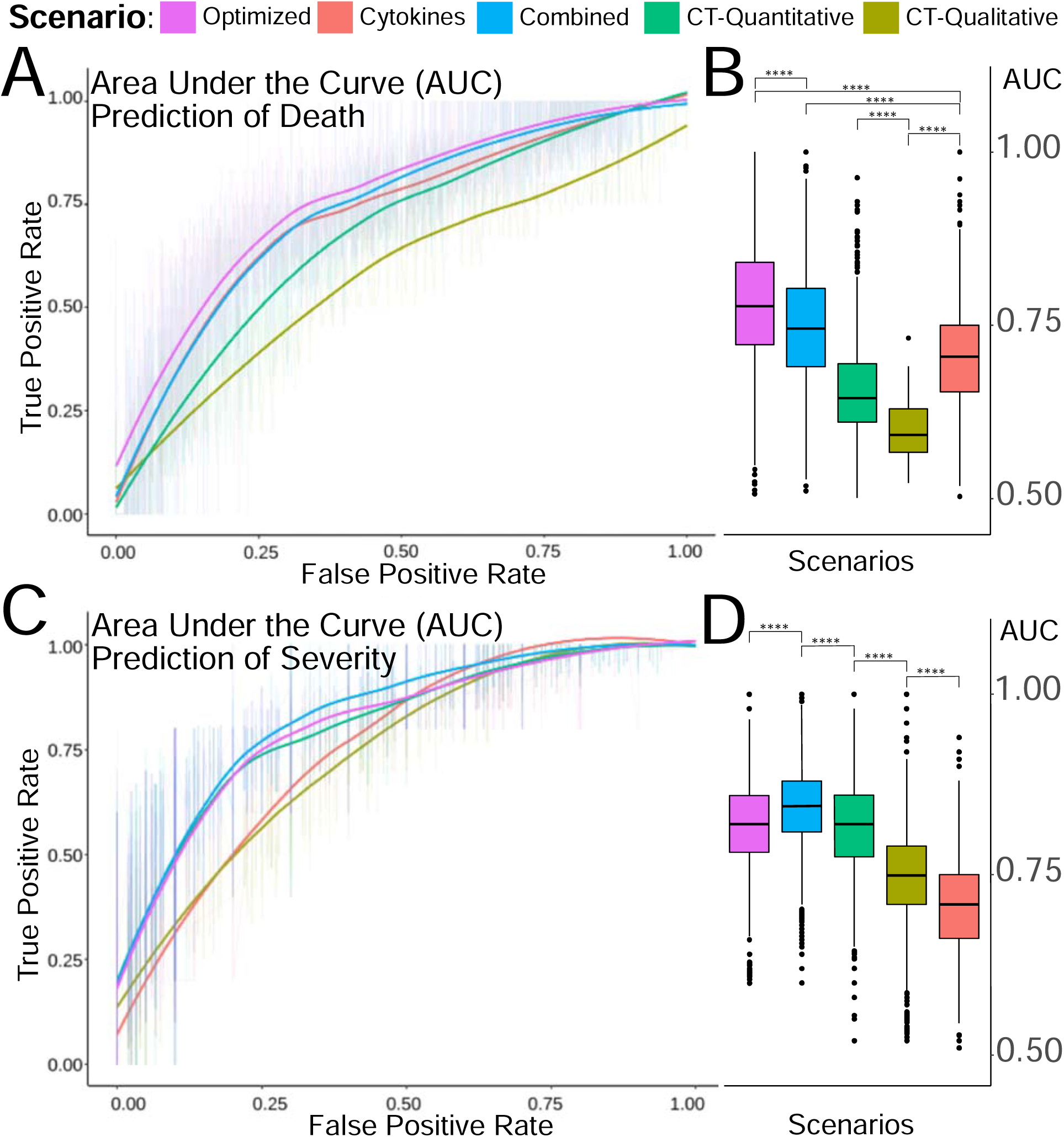
Power of chest CT and cytokines for prediction of death and maximum severity score. We tested the 5 scenarios to evaluate their relevance for prediction of death and maximum severity. A) Average ROC curves derived using risk of death are showed for each scenario. B) Boxplot showing the AUC values for all significant (p-value<0.05) models build per scenario for risk of death. C) Average ROC curves derived for maximum severity per scenario. D) Boxplot showing the AUC values for all significant (p-value<0.05) models build per scenario for severity.

The optimized prediction scenario contained IL-6, IL-8, TNF-α, GGO to aerated lung ratio, and age. This model showed an AUC of ~0.78 with CI 95% (72-84), combining cytokines (IL-6, IL-8, TNF-α,) and CT quantitative measurements (GGO to aerated lung ratio). The optimized scenario was significantly higher (adjusted p-values<0.05) than previous models using Wilcoxon rank test (**Figure 4B, 4D**). ^20^

### Prediction of COVID-19 maximum severity score

To test the predictive power of these scenarios for maximum COVID-19 severity (according to the WHO scale), we used elastic net regression. These results show that the combined scenario (AUC: 0.84 with CI 95% [80-88]) performed better (adjusted p-values<0.05) than the optimized scenario (AUC: 0.82 with CI 95% [78-86]) (adjusted p-value<0.05), CT quantitative (AUC: 0.81 with CI 95% [77-86]), CT qualitative (AUC: 0.74 with CI 95% [70-78]) (Adjusted p-values<0.05) (**Figure 4C, 4D**) and cytokines (AUC: 0.70 with CI 95% [66-75]) (Adjusted p-values<0.05). Of note, the CT quantitative performed better (Adjusted p-values<0.05) than CT qualitative and cytokines (**Figure 4D**).

Finally, to simplify our findings we took advantage of our elastic net regression interpretability to distillate probabilistic model for scoring risk or nomogram. The nomogram uses the glmnet selected variables GGO to aerated lung ratio, age, TNF-α, IL-6 and IL-8 to provide a score for risk of death (**Suppl. Figure 11**)^18 20^.

## DISCUSSION

Previous studies predicting COVID-19 patient outcomes using clinical, laboratory, or radiologic findings have shown overly optimistic predictive performance, partially due to risk of bias^22^. Moreover, these models did not utilize internationally validated classifications, such as the WHO scale, MODS, APACHE II, or SOFA. Thus, accurate prognostic models aimed at predicting COVID-19 outcomes are necessary.

In our study, we evaluated the performance of chest CT features and plasma cytokines from plasma alone and in combination in predicting death and maximum severity degree of hospitalized COVID-19 patients. These predictive models are important, so as to inform physicians of potential outcomes and thus helping with risk stratification, clinical decision making and treatment options. While cytokine assessment was more useful in prediction of death, CT features showed higher predictive performance for COVID-19 disease severity. In addition, the CT quantitative method, using volumetric variables extracted by manual segmentation outperformed CT qualitative score based on the radiologists’ assessment. Our findings reveal that a combined model using a CT quantitative feature (GGO to aerated lung ratio), demographics (age), and serum cytokines (IL-6, IL-8 and TNF-α) represents an accurate non-invasive tool for predicting risk of death and severity degree in hospitalized COVID-19 patients.

We investigated the probability of death using demographics, plasma cytokines (IL-6, IL-8, TNF-α), and CT measurements by Cox proportional hazard models. Although demographics variables showed significance (adjusted p-value<0.05) in univariate and multivariate analysis, they failed to predict outcomes with more than

0.6 AUC using elastic net regression. Further, we found that the models based on a combination of cytokines (IL-6, IL-8, and TNF-α) and age had a fair prediction power for death (AUC of 0.70) and maximum COVID-19 severity degree (AUC of 0.70). These results are concordant with the study carried out by Del Valle et al.^20^ that showed IL-6, IL-8, and TNF-α to be strong and independent predictors of patient death.

We assessed two approaches to evaluate CT images: one based on the radiologist’s qualitative assessment and a quantitative measurement of lesion burden using manual segmentation based on HU thresholding. Previous studies using either qualitative or quantitative image analysis methods have shown high performance in predicting adverse outcomes, such as mechanical ventilation, ICU admission, death, or severity degree ^7–11,13–17,35^. However, several of these studies used non-internationally validated severity degree scales.

Our models based on the CT qualitative and quantitative methods had an AUC of 0.61/0.66 for predicting death, respectively, underperforming when compared to cytokines (AUC 0.71). However, both CT models outperformed the cytokines model in predicting maximum severity score, with the CT quantitative model performing better than the CT qualitative model (AUCs of 0.81 and 0.74, respectively). These findings reveal that a quantitative assessment may more accurately assess lung involvement as stated by Avila et al.^6^. A model based on CT analyses could be a potential approach for COVID-19 stratification. In addition, both CT evaluation methods showed excellent inter-observer agreement between two independent readers with ICC > 0.9 in our study, concordant with other studies^8,11,16^

The potential of combining CT features with clinical and laboratory data (CRP, lymphocyte count and lymphocyte to neutrophil ratio among others) has been previously described^9,13,15,18^. However, the combination of CT features with plasma cytokines has not been evaluated. Our combined and optimized models, using both plasma cytokines and CT features, showed the best performances in predicting death (AUC 0.74 - 0.78) and maximum severity degree (AUC of 0.82 - 0.84). This noninvasive approach may help clinicians in risk stratification and in choosing individualized therapeutic strategies.

Our study design prevents overly broad conclusions. First, the sample size is too small to control for all types of potential biases. Our limited sample size was dictated by our strict inclusion/exclusion criteria. Second, the CT quantitative analysis could overestimate the GGO volume and consolidation volume due to partial volume effect and the inclusion of small vascular structures. However, this error affects every study and could be considered a systematic error that does not affect the final output. Finally, our population consists mainly of moderate to severe cases, as mild cases were not admitted.

Our study demonstrate that a combination of chest CT imaging analysis and plasma cytokine assessment can be used to profile COVID-19 patients and potentially triage them into risk groups. Here we show this combination to be a powerful tool in predicting COVID-19 mortality and severity degree. More importantly, we show that robust and simple predictive models can support clinical decision-making to stratify patients based on risk and aid in individualized therapeutic strategy development. Plasma cytokine assessment alone may help to predict survival, while CT analysis may aid in patient severity degree stratification. Additionally, CT quantitative analysis represents a potential tool to evaluate pneumonia lesions. Combining selected cytokine and CT quantitative features in an optimized model was shown to outperform either measurement alone.

Ultimately, these data and methods provide novel metrics that may be used to monitor patients upon hospitalization and help physicians make critical decisions and considerations for patients at high risk of death for COVID-19. These markers should be prospectively analyzed in relation to therapy choice, in particular with costly treatments such as monoclonal antibodies that could potentially be aimed at those patients with unfavorable scores at presentation.

## Supporting information

Main table 2

Main table 1

## Data Availability

The data supporting this publication has been made available at ImmPort (https://www.immport.org) under study accession number SDY1752. The dataset has been de-identified in compliance with United States Federal Health Insurance Portability and Accountability Act (HIPAA). ImmPort is a data sharing and analysis portal for immunology research community funded by the National Institute of Allergy and Infectious Diseases (NIAID), Division of Allergy, Immunology, and Transplantation (DAIT). Please refer to the ImmPort user agreement for further details (https://www.immport.org/agreement).

https://github.com/eegk/covid19_radiology

## ACKNOWLEDGEMENTS

Authors wish to acknowledge Rajiv Pande and Martin Putnam at Bio-techne for helping to provide instruments and assay kits for ELLA testing in a CLIA environment in the timeliest possible way during the health crisis. Additionally, this work was supported in part through the computational resources and staff expertise provided by Scientific Computing at the Icahn School of Medicine at Mount Sinai.

S.G., E.G.K, D.M.D.V. and M.M were supported by NCI U24 grant CA224319. S.G. and D.M.D.V. is additionally supported by grant U01 DK124165.

## ABBREVIATIONS

AUC: Area Under the Curve
COVID-19: Novel coronavirus discovered in 2021 Wuhan
CT: Computed Tomography
EMR: Electronic Medical records
GGO: Ground-Glass Opacities
HU: Hounsfield Units
IL-6: Interleukin 6
IL-8: Interleukin 8
IQR: Inter Quartile Range
ROC: Receiver Operating Characteristic
TNF-α: Tumor Necrosis Factor alpha
WHO: World Health Organization

## REFERENCES

1. Chen, N., et al. Epidemiological and clinical characteristics of 99 cases of 2019 novel coronavirus pneumonia in Wuhan, China: a descriptive study. The Lancet 395, 507–513 (2020).

2. Phillips, N. The coronavirus is here to stay-here’s what that means. Nature 590, 382–384 (2021).

3. Bernheim, A., et al. Chest CT findings in coronavirus disease-19 (COVID-19): relationship to duration of infection. Radiology, 200463 (2020).

4. Shi, H., et al. Radiological findings from 81 patients with COVID-19 pneumonia in Wuhan, China: a descriptive study. The Lancet infectious diseases 20, 425–434 (2020).

5. Mei, X., et al. Artificial intelligence-enabled rapid diagnosis of patients with COVID-19. Nat Med 26, 1224–1228 (2020).

6. Avila, R.S., et al. QIBA guidance: Computed tomography imaging for COVID-19 quantitative imaging applications. Clinical imaging (2021).

7. Colombi, D., et al. Well-aerated lung on admitting chest CT to predict adverse outcome in COVID-19 pneumonia. Radiology, 201433 (2020).

8. Colombi, D., et al. Qualitative and quantitative chest CT parameters as predictors of specific mortality in COVID-19 patients. Emergency radiology 27, 701–710 (2020).

9. Feng, Z., et al. Early prediction of disease progression in COVID-19 pneumonia patients with chest CT and clinical characteristics. Nature communications 11, 1–9 (2020).

10. Francone, M., et al. Chest CT score in COVID-19 patients: correlation with disease severity and short-term prognosis. European Radiology, 1–10 (2020).

11. Li, K., et al. CT image visual quantitative evaluation and clinical classification of coronavirus disease (COVID-19). European radiology, 1–10 (2020).

12. Lyu, P., Liu, X., Zhang, R., Shi, L. & Gao, J. The performance of chest CT in evaluating the clinical severity of COVID-19 pneumonia: identifying critical cases based on CT characteristics. Investigative Radiology 55, 412–421 (2020).

13. Matos, J., et al. Evaluation of novel coronavirus disease (COVID-19) using quantitative lung CT and clinical data: prediction of short-term outcome. European radiology experimental 4, 1–10 (2020).

14. Park, B., et al. Prognostic implication of volumetric quantitative ct analysis in patients with COVID-19: a multicenter study in Daegu, Korea. Korean journal of radiology 21, 1256 (2020).

15. Sun, D., et al. CT Quantitative Analysis and Its Relationship with Clinical Features for Assessing the Severity of Patients with COVID-19. Korean Journal of Radiology 21, 859–868 (2020).

16. Yang, R., et al. Chest CT severity score: an imaging tool for assessing severe COVID-19. Radiology: Cardiothoracic Imaging 2, e200047 (2020).

17. Yin, X., et al. Assessment of the severity of coronavirus disease: quantitative computed tomography parameters versus semiquantitative visual score. Korean journal of radiology 21, 998 (2020).

18. Zheng, Y., et al. Development and validation of a prognostic nomogram based on clinical and CT features for adverse outcome prediction in patients with COVID-19. Korean journal of radiology 21, 1007 (2020).

19. Mehta, P., et al. COVID-19: consider cytokine storm syndromes and immunosuppression. The lancet 395, 1033–1034 (2020).

20. Del Valle, D.M., et al. An inflammatory cytokine signature predicts COVID-19 severity and survival. Nature medicine 26, 1636–1643 (2020).

21. Ragab, D., Salah Eldin, H., Taeimah, M., Khattab, R. & Salem, R. The COVID-19 cytokine storm; what we know so far. Frontiers in immunology 11, 1446 (2020).

22. Wynants, L., et al. Prediction models for diagnosis and prognosis of covid-19: systematic review and critical appraisal. bmj 369(2020).

23. Chung, M., et al. CT imaging features of 2019 novel coronavirus (2019-nCoV). Radiology 295, 202–207 (2020).

24. Hansell, D.M., et al. Fleischner Society: glossary of terms for thoracic imaging. Radiology 246, 697–722 (2008).

25. Kikinis, R., Pieper, S.D. & Vosburgh, K.G. 3D Slicer: a platform for subject-specific image analysis, visualization, and clinical support. in Intraoperative imaging and image-guided therapy 277–289 (Springer, 2014).

26. World Health Organization. COVID-19 Therapeutic Trial Synopsis. (2020).

27. Bonett, D.G. & Wright, T.A. Sample size requirements for estimating Pearson, Kendall and Spearman correlations. Psychometrika 65, 23–28 (2000).

28. Bower, K.M. When to use Fisher’s exact test. vol. 2 35–37.

29. Core Team, R. R: A language and environment for statistical computing. R Foundation for Statistical Computing. Vienna, Austria: URL https://www.R-project.org/.[Google Scholar] (2017).

30. Friedman, J., Hastie, T. & Tibshirani, R. Regularization paths for generalized linear models via coordinate descent. Journal of statistical software 33, 1 (2010).

31. Baldi, P., Brunak, S. & Bach, F. Bioinformatics: the machine learning approach, (MIT press, 2001).

32. Robin, X., et al. pROC: an open-source package for R and S+ to analyze and compare ROC curves. BMC bioinformatics 12, 1–8 (2011).

33. Harrell, F.E. RMS: regression modeling strategies. R package version 51-2, 2018. Точкадоступа: http://CRAN.R-project.org (2019).

34. Benchoufi, M., Matzner-Lober, E., Molinari, N., Jannot, A.S. & Soyer, P. Interobserver agreement issues in radiology. Diagnostic and Interventional Imaging 101, 639–641 (2020).

35. Lanza, E., et al. Quantitative Chest CT analysis in COVID-19 to predict the need for oxygenation support and intubation. (2020).

