## Supplementary material for "Quantitative chest CT combined with plasma cytokines predict outcomes in COVID-19 patients": Suplemental legends and text

**Supplementary Figure Legends**

**Supplementary Figure 1.** Spearman correlation heatmap showing colored circles (red for positive correlation and blue for negative correlation) for each comparison between all numeric variables used in this study. Additionally, the heatmap is hierarchically clustered showing associations between variables that are close to each. Immediately visible is two main clusters, one for chest CT measurements and one for cytokines.

**Supplementary Figure 2**. Cox proportional hazard model showing that age separated in 4 quantiles does not show significant differences for the probability of survival.

**Supplementary Figure 3**. Cox proportional hazard model showing that gender separated in male and female does not show significant differences for the probability of survival.

**Supplementary Figure 4**. Cox proportional hazard model showing that BMI separated in 4 quantiles does not show significant differences for the probability of survival.

**Supplementary Figure 5**. Cox proportional hazard model showing that O2 saturation separated in 4 quantiles showed significant differences for the probability of survival.

**Supplementary Figure 6**. Cox proportional hazard model showing that IL-6 separated in low/high show significant differences for the probability of survival.

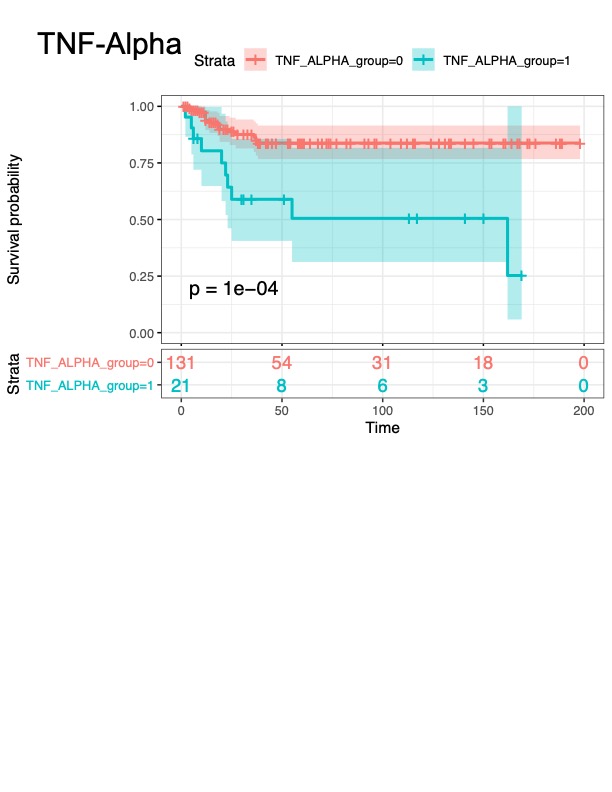

**Supplementary Figure 7**. Cox proportional hazard model showing that TNF-α separated in low/high show significant differences for the probability of survival.

**Supplementary Figure 8**. Cox proportional hazard model showing that IL-8 separated in low/high show significant differences for the probability of survival.

**Supplementary Figure 9**. Cox proportional hazard model showing that CT qualitative score separated in quantiles showed not significant differences for the probability of survival.

**Supplementary Figure 10**. Cox proportional hazard model showing that GGO Aerated lung ratio score separated in quantiles showed significant differences for the probability of survival.

**Supplementary Figure 11**. Risk of death Nomogram for covid19 build using the 5 key variables chosen by elastic net coefficient selection including: GGO to aerated lung ratio, age, TNF-α, IL6 and IL8. The measurements are binarized for TNF-α, IL-6 and IL-8. Once each variable is scored, the points attributed are located on top of the nomogram. Then, all points are summed and compared to the points scale on the bottom of the figure, where risk of death is attributed based on this sum.

**Supplementary table 1.** CT vendors and acquisition parameters for each scanner. ST: slice thickness; PS: pixel spacing.

**Supplementary table 2.** CT qualitative variables described by radiologists according to the Fleischner Society guidelines.

**Supplementary table 3.** Statistical tests comparing available the average values for variables between patients that die or survive.

**Supplementary table 4.** Univariate Survival Cox proportional hazard models for all available variables separated in classes or 4 quartiles depending on the variable.

**Supplementary table 5.** Multivariate Survival Cox proportional hazard models for relevant variables selected as important after correlation and coxph univariate analysis.

**Supplementary table 6.** Elastic net regression summary for patients that died and those who didn’t for models using Oxygenation and demographics. The column alpha indicates the mixing coefficient for lasso and ridge proportion. Prob. indicates the percentage of samples used as testing and AUC indicates the area under the curve values.
